# Proton Therapy Reduces Cardiopulmonary Dose in Synchronous Bilateral Breast Cancer, Especially with Direct Chest Wall Irradiation

**DOI:** 10.1101/2025.08.01.25332746

**Authors:** Noah Khosrowzadeh, Kyle Chambers, Aren Singh Saini, Kayla Samimi, Matthew Gompels, Cyrus Washington, Jessica Meshman, Cristiane Takita

## Abstract

**Purpose/Objective(s):** To evaluate the dosimetric advantages of intensity-modulated proton therapy (IMPT) compared to volumetric modulated arc therapy (VMAT) in patients with synchronous bilateral breast cancer (sBBC), with a focus on cardiopulmonary sparing, particularly in patients receiving direct chest wall irradiation.

**Methods and Materials:** A retrospective dosimetric analysis was conducted on 11 patients with sBBC treated with both IMPT and VMAT plans between 2020 and 2021 at a tertiary cancer center. Target volumes included bilateral breasts/chest wall and nodal regions. Mean dose V5, and V10, values were compared for the heart, lungs, and left anterior descending artery (LAD) using Wilcoxon signed-rank tests. V20 was compared for the heart and lungs. A subanalysis was performed for patients who received direct chest wall radiation.

**Results:** IMPT significantly reduced mean heart dose (−522 cGy, *p* = 0.0034), heart V5 (−67.77%, *p* = 0.0076), and heart V10 (−0.91%, *p* = 0.0164). Lung exposure was also lower with IMPT: mean dose (−988 cGy, *p* = 0.0034), V5 (−52.38%, *p* = 0.0044), V10 (−30.21%, *p* = 0.0044), and V20 (−19.68%, *p* = 0.0034). When looking at the left anterior descending artery (LAD), IMPT significantly reduced mean LAD dose (−509 cGy, *p* = 0.0034), V5 (−88.12%, *p* = 0.0069), and V10 (−11.47%, *p* = 0.0093). In the chest wall subcohort, IMPT showed greater sparing of heart (−792 cGy), lungs (−820 cGy), and LAD (−1117 cGy), all with statistically significant *Z*-scores.

**Conclusion:** IMPT significantly reduces radiation dose to the heart, lungs, and LAD in patients with sBBC compared to VMAT, particularly in those receiving direct chest wall irradiation. These findings support expanding access to proton therapy and suggest that current insurance approval criteria should include chest wall radiation as an indication.

## Introduction

Breast cancer is the most commonly diagnosed cancer among women and the fourth leading cause of cancer-related death worldwide [1]. Synchronous bilateral breast cancer (sBBC) is a rare subtype of breast cancer, accounting for 2.9-3.9% of all breast cancer diagnoses [2]. sBBC is defined as breast cancer identified in each breast at the same time. Risk factors for sBBC include family history of breast cancer, younger age at diagnosis, and lobular histology. Overall, breast cancer incidence and sBBC incidence have both increased in the United States according to the Surveillance, Epidemiology, and End Results (SEER) database of the National Cancer Institute [3]. Patients with sBBC tend to have worse overall survival and recurrence-free survival rates compared to those with unilateral breast cancer [4–6].

The current standard of care for breast cancer involves a multidisciplinary approach combining surgery, radiotherapy, and other neoadjuvant and adjuvant therapies [5]. However, there is no established standard of care in terms of radiotherapy for patients with sBBC due to the technical complexity of treatment. Many patients with sBBC will require bilateral breast irradiation. This type of radiotherapy necessitates larger target volumes, which can result in high radiation doses due to field overlap, insufficient lymph node coverage, and significant exposure to cardiopulmonary structures [7, 8].

The use of traditional three-dimensional conformal radiation therapy (3D-CRT) in bilateral breast irradiation results in midline beam overlap with constrained beam arrangement options, leading to increased radiation exposure to adjacent organs at risk. Volumetric modulated arc therapy (VMAT) and Intensity Modulated Radiation Therapy (IMRT) dosimetry has improved normal tissue sparing[8, 9]. Nevertheless, studies have shown that VMAT doses to the heart and lungs in sBBC frequently exceed standard dose constraints [10, 11].

The association between radiation exposure to the thoracic region and cardiovascular disease is well documented [12]. This risk is particularly pronounced during radiotherapy for bilateral breast or chest wall irradiation due to the elevated dose delivered to both the heart and lungs. Radiation-induced cardiac toxicity can manifest acutely as pericarditis, or chronically as coronary artery disease, myocardial infarction, or congestive heart failure [13–15]. Studies have reported a relative risk ranging from 1.2 to 1.3 for these conditions, with the risk increasing in proportion to the radiation dose received [13]. A linear dose–response relationship has been demonstrated, indicating a 7.4% increase in the incidence of ischemic heart disease per Gray (Gy) of mean heart dose (MHD) [14]. While MHD remains a critical parameter, high-dose regions often involve specific substructures, particularly the anterior heart and the left anterior descending (LAD) coronary artery [16,17]. Notably, the LAD has been shown to receive disproportionately higher doses relative to the remainder of the heart and is a key contributor to radiation-induced ischemic heart disease [16]. Similarly, pulmonary toxicity is a significant concern, with radiation exposure to the lungs associated with acute radiation pneumonitis and late-stage pulmonary fibrosis [18]. These toxicities correlate with both the mean lung dose and the volume of lung receiving ≥10, 20, 30, 40, or 50 Gy [19]. Therefore, meticulous attention to both cardiac and pulmonary dosimetry is essential to minimize treatment-related morbidity and improve clinical outcomes in breast cancer patients.

Intensity-modulated proton therapy (IMPT) has emerged as a promising alternative, demonstrating better organ at risk (OAR) sparing compared to photon therapies like VMAT [20, 21]. Even with photon optimization techniques like deep inspiration breath hold, proton therapy still outperformed photon therapy in sparing critical cardiac substructures, which is particularly important in breast cancer cases where breath-hold may not be feasible [22].

Previous studies comparing proton and photon therapy in unilateral breast cancer have demonstrated superior target coverage and reduced acute toxicity with intensity-modulated proton therapy (IMPT) [21,22]. However, the application of IMPT in synchronous bilateral breast cancer (sBBC) remains underexplored, with only three prior studies—each limited by small sample sizes—focusing on its potential for normal tissue sparing. These studies consistently reported improved sparing of the heart, lungs, and left anterior descending (LAD) artery, along with enhanced dose conformity to target volumes [23–25]. Given the growing incidence and complexity of managing sBBC, a thorough understanding of its radiation dosimetry is critical [26]. The present study aims to assess whether the dosimetric advantages of IMPT over volumetric modulated arc therapy (VMAT) are sufficiently robust to support its use as the preferred radiotherapy modality in this high-risk patient population. To this end, a dosimetric comparison between IMPT and VMAT was conducted in patients with sBBC, with particular attention to doses delivered to the heart, lungs, and LAD.

In addition, current indications for primary insurance approval for proton therapy as opposed to photon therapy as the standard of care do not include direct chest wall radiation and insurers typically require evidence that photon therapy cannot meet dose constraints to organs at risk (OARs) before approving proton therapy. However, there remains the room for investigation that direct chest wall radiation with photon therapy may demonstrate worse outcomes for these patients due to the proximity of the target to vital structures such as the heart, coronary arteries, and lungs.

## Materials/Methods

A retrospective review was conducted of patients with synchronous bilateral breast cancer who were treated with intensity-modulated proton therapy (IMPT) between September 2020 and February 2021 at a tertiary cancer center. Contouring plans were created during the initial CT simulation by physicists [Figure 1]. The clinical target volume (CTV) was contoured with reference to the Radiation Therapy Oncology Group contouring atlas and was defined as the volume of the bilateral breast, chest walls and bilateral axillary lymph nodes. IMPT prescriptions were for 45-50 Gy in 25 fractions with 95% of the target volumes covered by 95% of the prescription dose and dose constraints to OARs per published prospective trials. VMAT plans were designed for the same patients with the same target coverage and OAR constraints for comparison. For each patient, dosimetry data was collected from the theoretical proton and photon treatment plans using the ARIA Radiation Oncology Information (Varian Medical Systems). The mean dosages to target tissues were documented in cGy and compared to corresponding values from VMAT plans, which were created in collaboration with physicists and analyzed using Excel. The data was considered paired. The V5, V10, and V20 radiation doses delivered to the heart, lungs, and left anterior descending (LAD) artery were also documented for each treatment plan. Additionally, patients receiving direct chest wall radiation were isolated for further dosimetric analysis. For these patients, mean dose exposures specifically to the chest wall regions were compared between proton and photon plans. In total, 11 patients had paired dosimetry data available for the heart, lungs, and LAD. To determine the appropriate statistical test, the distribution of the dosages for proton and photon mean radiation was assessed using the Jarque-Bera test for normality. The distributions were found to be non-normal (p<0.05). Consequently, a Wilcoxon signed-rank test was used to compare organ dose exposures between the paired proton and photon plans. Statistical significance was defined as <0.05. Analyses were performed using Microsoft Excel.

**Figure 1.**
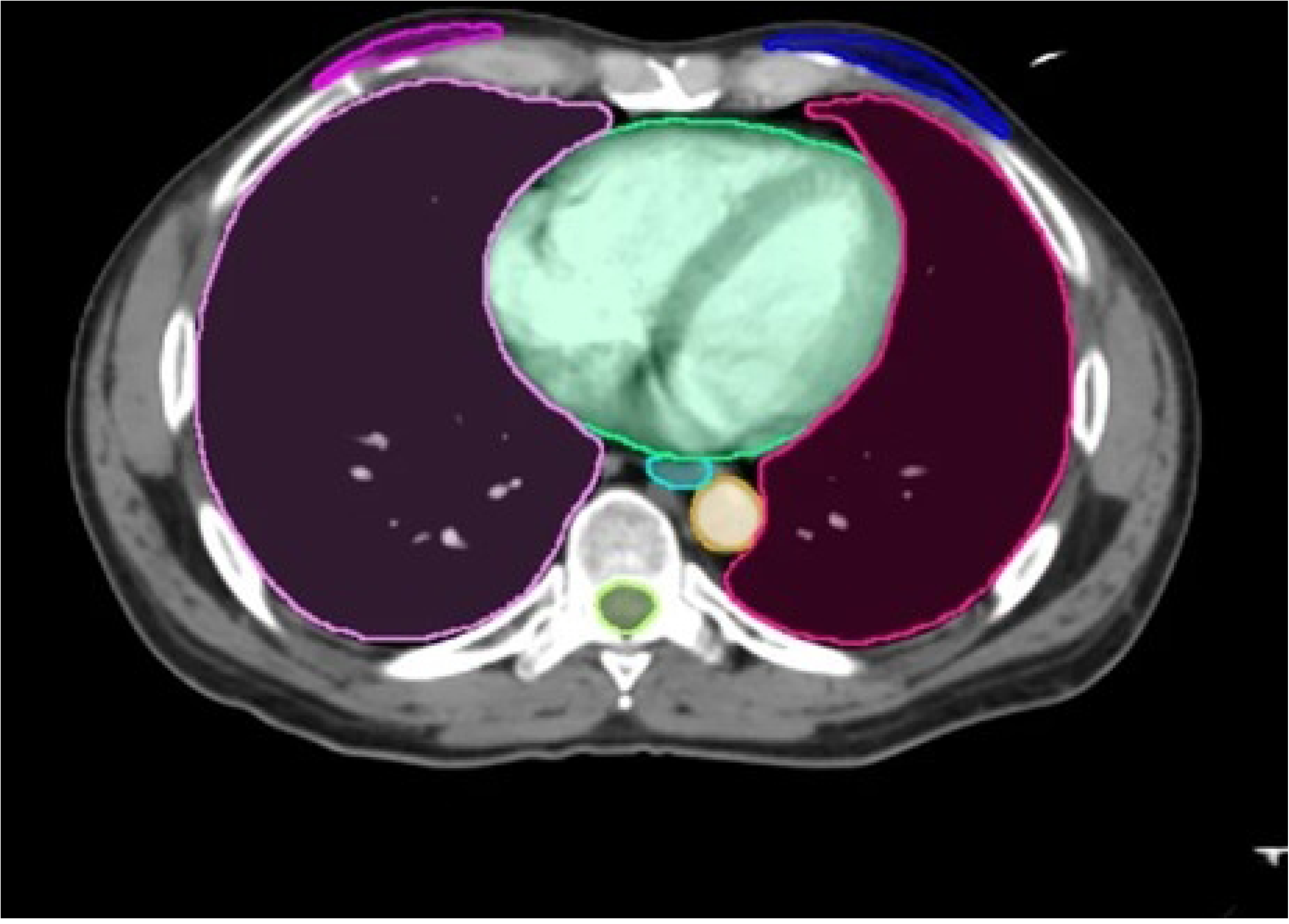
Organs at Risk Contouring CT Scan. An example of the heart, lungs, and other structure contours overlaid on a treatment planning CT scan. These structures, including the LAD (which is not contoured in this example), were used to extract mean dose and volume-based metrics (V5, V10, V20) for statistical comparison in our study.

This study was approved by the Institutional Review Board (IRB) at the University of Miami Miller School of Medicine (IRB approval number: # 20090856). Due to the retrospective nature of this dosimetric analysis, the IRB waived the requirement for informed consent, as all patient data were fully anonymized and analyzed in aggregate without identifiable personal information.

## Results

A total of 11 patients had paired dosimetry data available for both proton and photon treatment plans. To compare the radiation exposure between modalities for each organ at risk, a Wilcoxon signed-rank test was conducted. For heart, lungs, and LAD mean dosage in cGy was compared [Table 1, Figure 2, Figure 3] as well as percentage volume metrics of V5 and V10 [Figure 4, Figure 5, Figure 6]. V20 percentage volume metrics were compared for heart and lungs [Figure 4, Figure 5].

**Figure 2.**
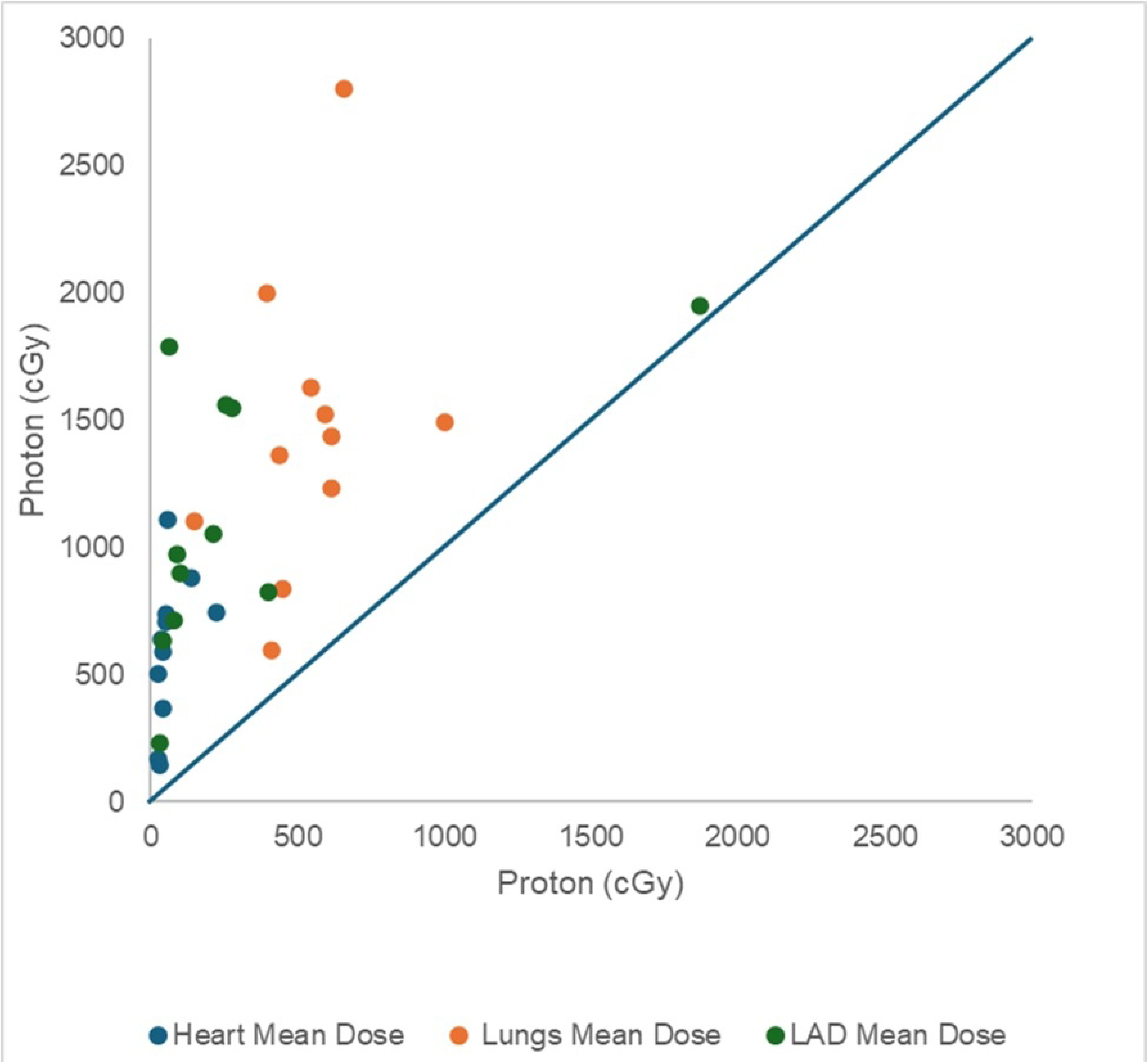
Comparison of Heart, Lungs, and LAD mean dose exposure between competing proton and photon plans for each patient. Each point represents a patient’s mean dose to a given organ. The diagonal line (y = x) indicates equal dosing. Nearly all points fall above the line, demonstrating that photon therapy consistently delivered higher mean doses to the heart, lungs, and LAD compared to proton therapy.

**Figure 3.**
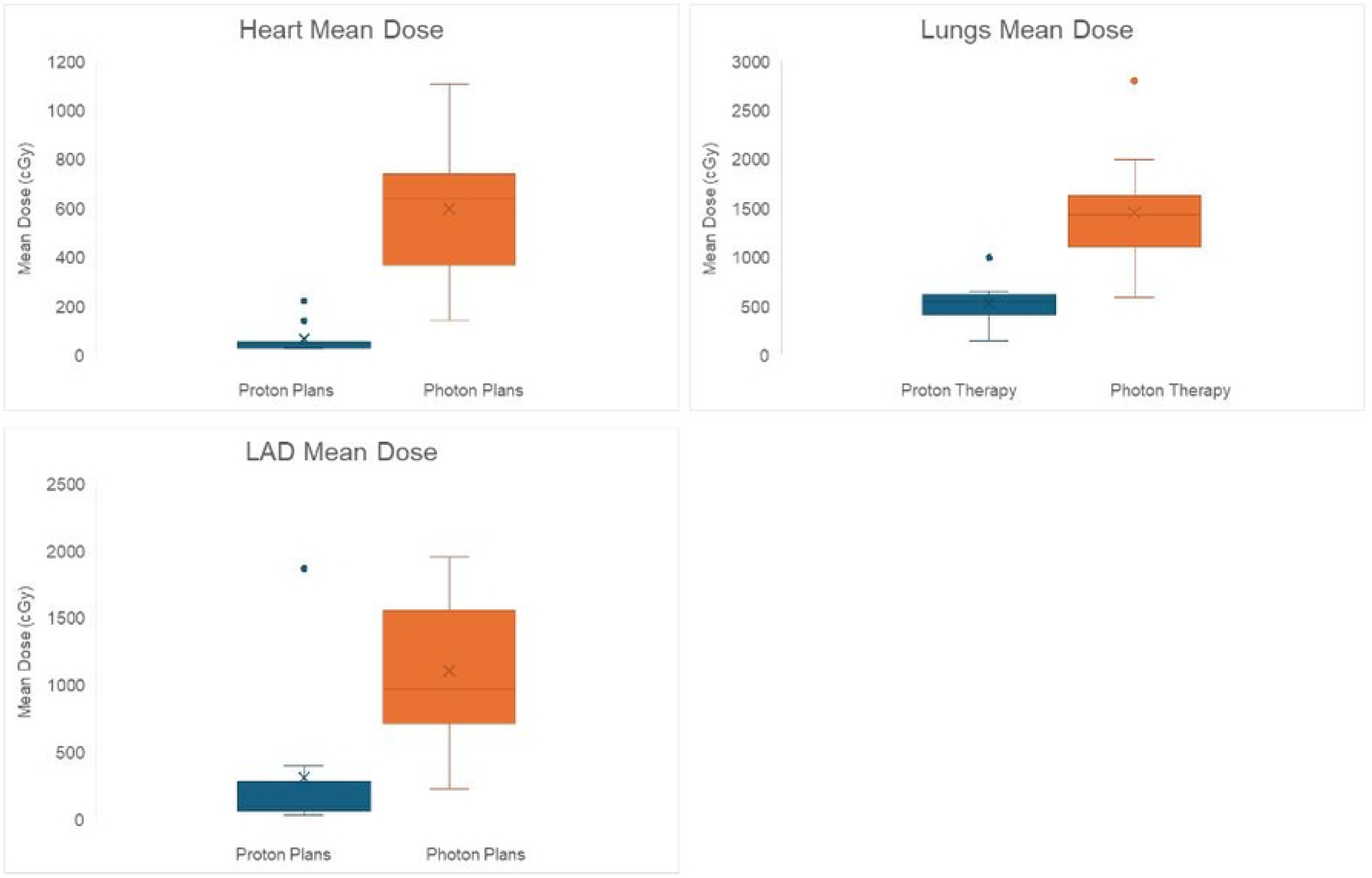
Heart, Lung, and LAD Mean Dose. Box plots demonstrating the mean radiation doses (cGy) delivered to the heart (A), lungs (B), and LAD artery (C). Proton therapy plans delivered significantly lower mean doses compared to photon therapy plans across all three organs.

**Figure 4.**
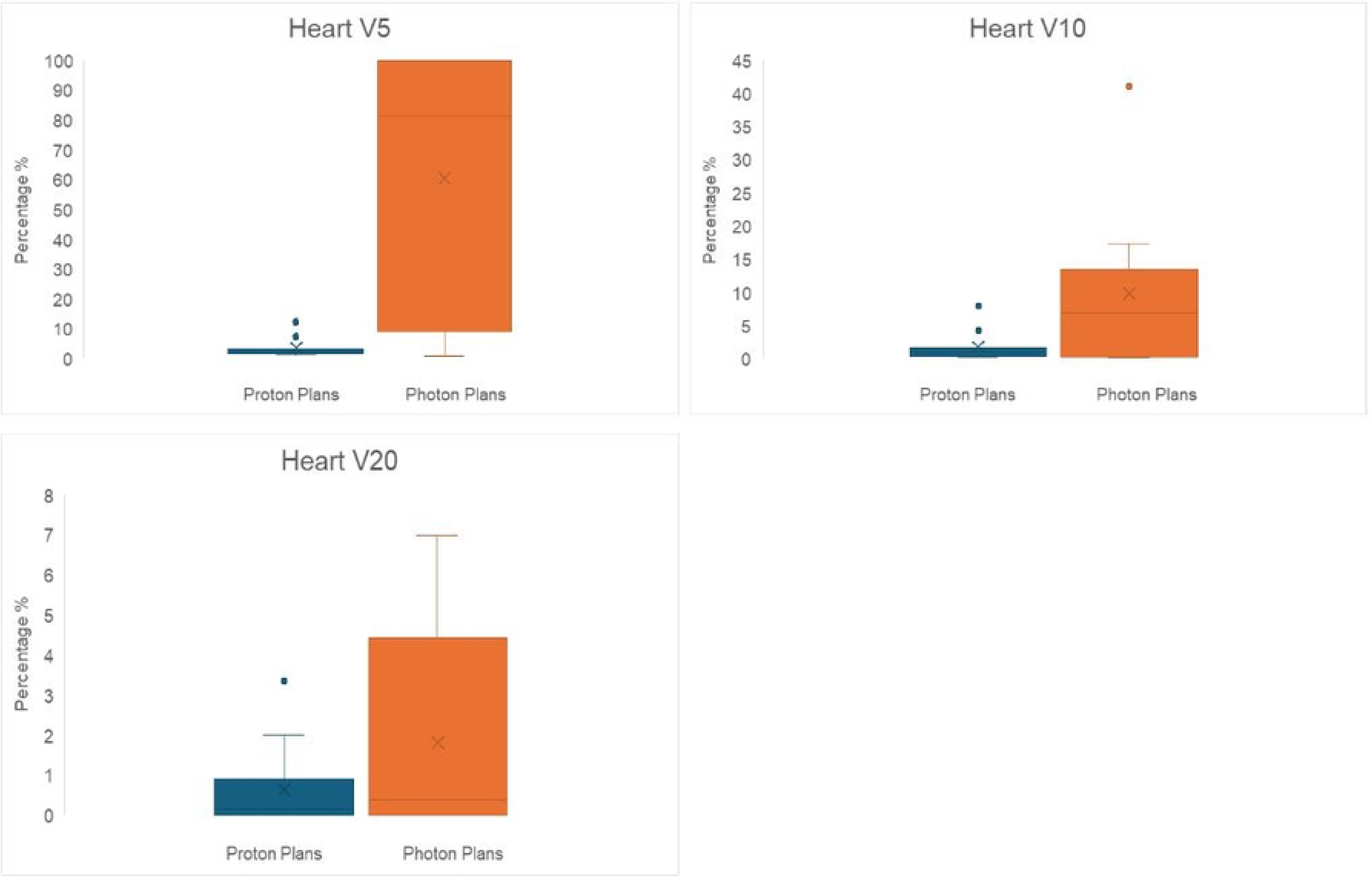
V5, V10, V20 of the Heart. Box plots showing the percentage of the heart receiving at least 5 Gy (V5) [A], 10 Gy (V10) [B], and 20 Gy (V20) [C]. Proton therapy plans consistently reduced low-, intermediate-, and high-dose heart exposures compared to photon therapy.

**Figure 5.**
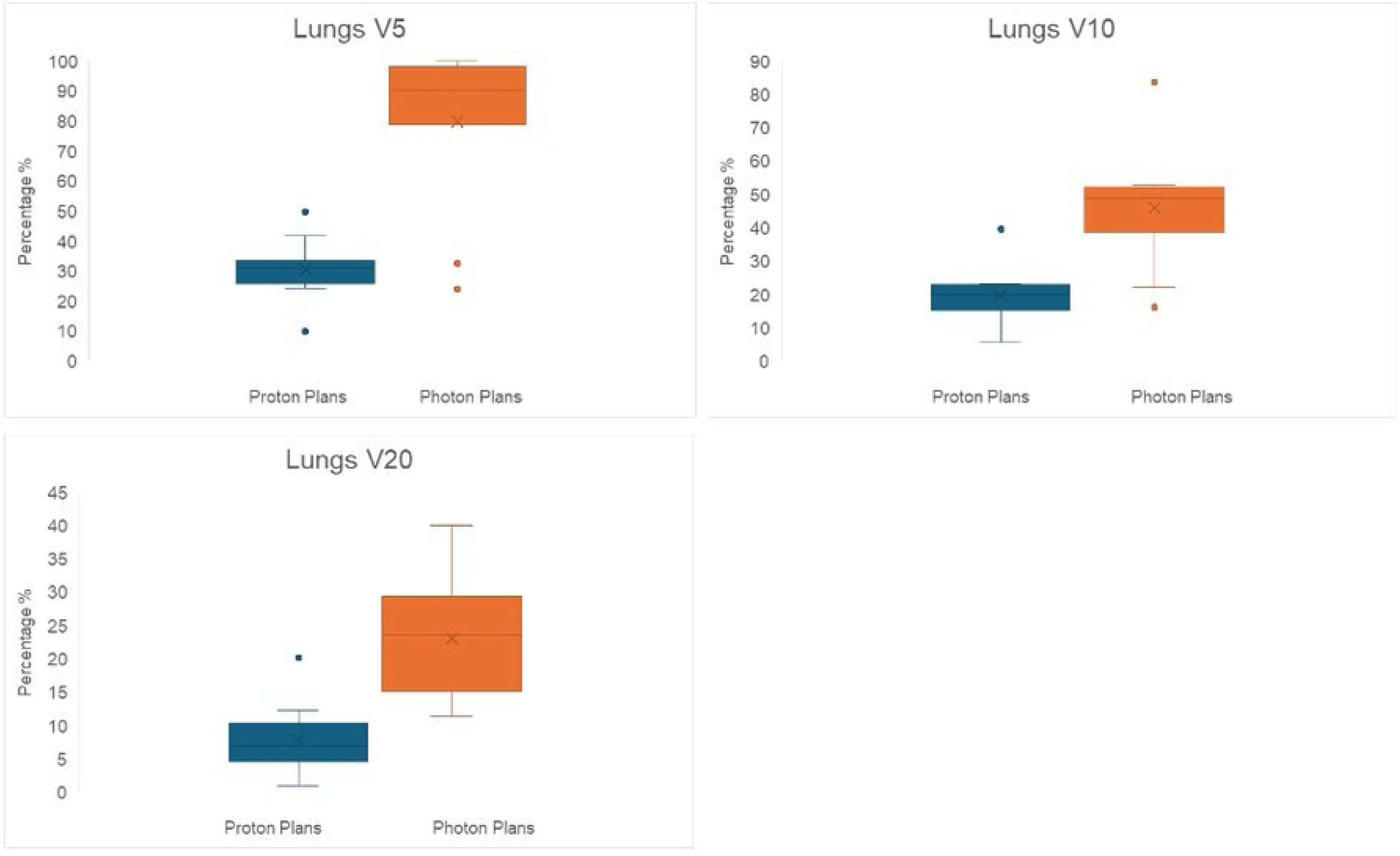
V5, V10, V20 of Lungs. Box plots comparing the percentage of lung volume receiving at least 5 Gy (V5) [A], 10 Gy (V10) [B], and 20 Gy (V20) [C]. Proton therapy demonstrated reduced radiation exposure to lung tissue compared to photon therapy across all dose levels.

**Figure 6.**
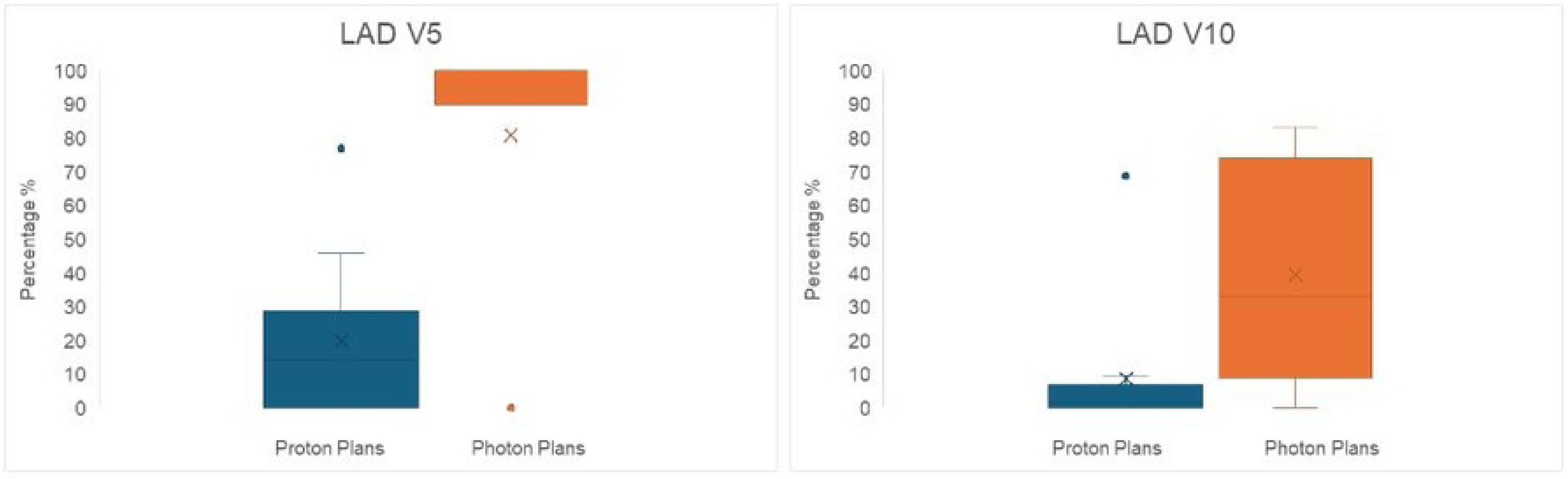
LAD V5 and V10. Box plots illustrating the percentage of the LAD artery receiving at least 5 Gy (V5) [A] and 10 Gy (V10) [B]. Proton therapy markedly reduced LAD artery radiation doses compared to photon therapy.

**Figure 7.**
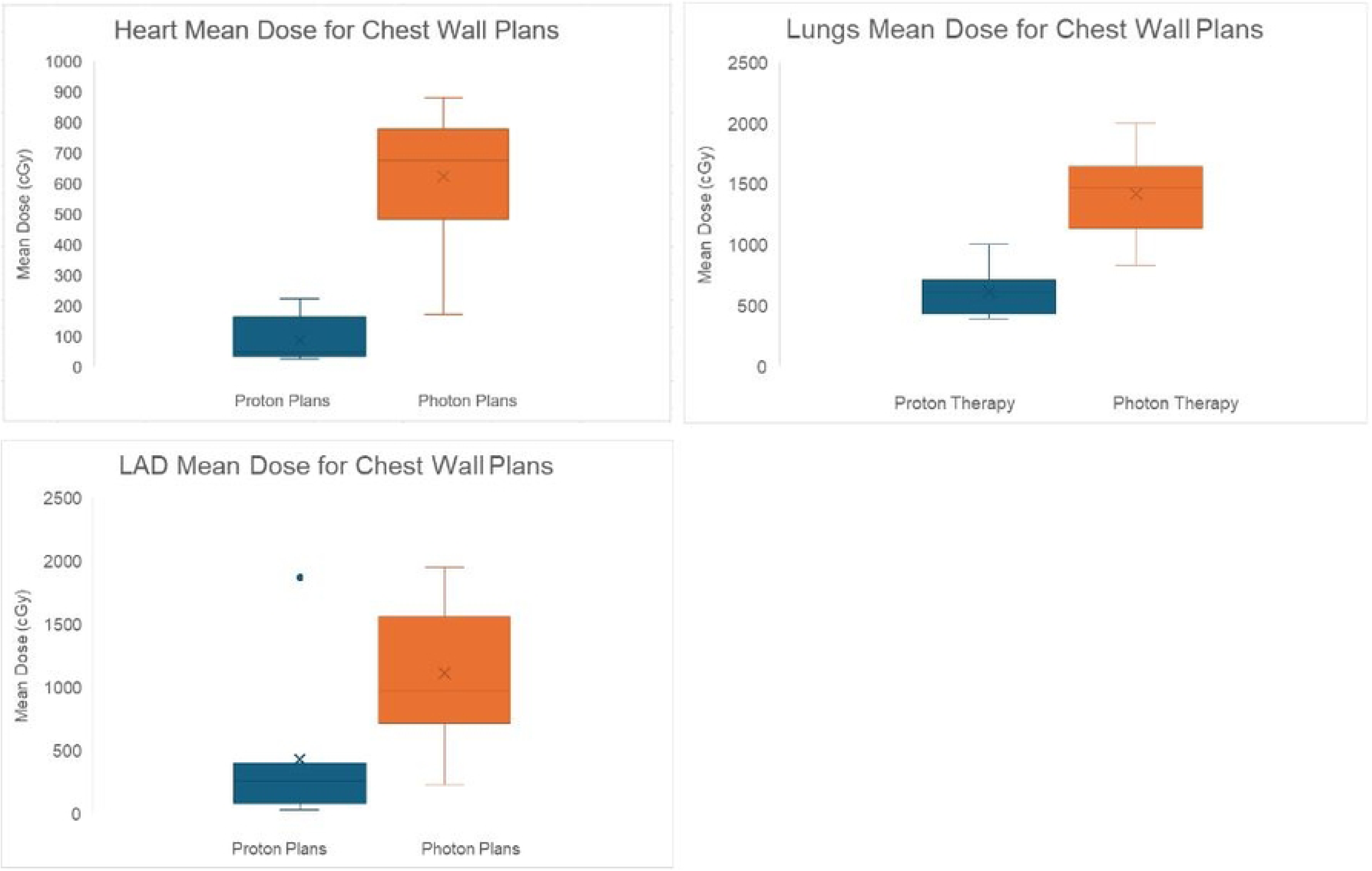
Heart, Lung, and LAD Mean Dose for Chest Wall Plans. Box plots demonstrating the mean radiation doses (cGy) delivered to the heart (A), lungs (B), and LAD artery in direct chest wall radiation plans. Proton therapy plans delivered significantly lower mean doses compared to photon therapy plans across all three organs.

**Table 1.** Organs at Risk Dosage Differences. Summary of mean dosimetric differences between proton therapy and photon therapy plans across organs at risk, including the heart, lungs, and left anterior descending (LAD) artery.

| Organ | Parameter | Median Difference<br>(cGy or %) | W-value | Z-value | p-value | Significant |
| --- | --- | --- | --- | --- | --- | --- |
| Heart | Mean Dose<br>(cGy) | -521.65 | 0 | -2.93 | 0.0034 | Yes |
|  | V5 (%) | -67.77 | 3 | -2.67 | 0.0076 | Yes |
|  | V10 (%) | -0.91 | 6 | -2.40 | 0.0164 | Yes |
|  | V20 (%) | +0.62 | 11 | -1.96 | 0.0500 | No |
| Lungs | Mean Dose<br>(cGy) | -988.25 | 0 | -2.93 | 0.0034 | Yes |
|  | V5 (%) | -52.38 | 1 | -2.85 | 0.0044 | Yes |
|  | V10 (%) | -30.21 | 1 | -2.85 | 0.0044 | Yes |
|  | V20 (%) | -19.68 | 0 | -2.93 | 0.0034 | Yes |
| LAD | Mean Dose<br>(cGy) | -509.17 | 0 | -2.93 | 0.0034 | Yes |
|  | V5 (%) | -88.12 | 1 | -2.70 | 0.0069 | Yes |
|  | V10 (%) | -11.47 | 2 | -2.59 | 0.0093 | Yes |

### Heart dosage comparison

In terms of radiation exposure to the heart, proton therapy plans resulted in a significantly lower mean heart dose compared to that of photon plans [Table 1, Figure 2A]. The median difference was determined to be -521.65 cGy, with all patients in the data set exhibiting reduced heart exposure dose under proton therapy (W = 0, Z = −2.93, p = 0.0034; sum of negative ranks = 66) [Table 1]. For the V5 measurement of the heart, proton therapy also elicited significantly reduced exposure with a median difference of -67.77% with 10 of 11 patients showing reduced values (W = 3, Z = -2.67, p = 0.0076, sum of negative ranks = 63) [Table 1, Figure 4A]. Heart V10 measurement again showed significantly lower exposure with proton therapy plans compared to photon plans with a median difference of -0.91% (W =6, Z = -2.93, p = 0.0164, sum of negative ranks = 60) [Table 1, Figure 4B]. For heart V20, the median difference between proton and photon therapy plans was determined to be +0.62%. While 10 of the 11 patients had a lower V20 with proton therapy, this result narrowly missed statistical significance (W = 11, Z = -1.96, p = 0.05, sum of negative ranks = 55) [Table 1, Figure 4C].

### Lung dosage comparison

In terms of lung exposure, results similarly echoed that of the heart with a significantly reduced mean lung dose. The median difference was -988.25 cGy with all patients experiencing lower lung exposure with proton therapy (W = 0, Z = -2.93, p = 0.0034, sum of negative ranks = 66) [Table 1, Figure 2B]. Proton therapy was also found to have significantly lower lung V5 values with a median difference of -52.38%. 10 of the 11 patients showed reduced values for proton therapy plans (W = 1, Z = -2.85, p = 0.0044, sum of negative ranks = 65) [Table 1, Figure 5A]. V10 values also mirrored V5 with a median difference of -30.21% (W = 1, Z = -2.85, p = 0.0044, sum of negative ranks = 65) [Table 1, Figure 5B]. For V20, proton therapy was determined to have a median difference of -19.68%. All patients had lower V20 values for proton therapy when compared to photon therapy plans (W = 0, Z = -2.93, p = 0.0034, sum of negative ranks = 66) [Table 1, Figure 5C].

### LAD dosage comparison

For radiation exposure to the LAD, proton therapy plans resulted in a significantly lower mean LAD dose compared to that of photon plans [Table 1, Figure 2C]. The median difference was determined to be -509.17 cGy, with all patients in the data set exhibiting reduced LAD exposure dose under proton therapy (W = 0, Z = −2.93, p = 0.0034; sum of negative ranks = 66) [Table 1]. For the V5 measurement of the LAD, proton therapy also elicited significantly reduced exposure with a median difference of -88.12% with 10 of 11 patients showing reduced values (W = 1, Z = -2.7011, p = 0.0069, sum of negative ranks = 54) [Table 1, Figure 6]. LAD V10 measurement again showed significantly lower exposure with proton therapy plans compared to photon plans with a median difference of -11.47% (W = 2, Z = -2.59, p = 0.0093, sum of negative ranks = 53) [Table 1, Figure 6]. Analysis of V20 was not conducted as there was an insufficient number of patients required that met this threshold.

### Direct chest wall radiation exposure

In patients receiving direct chest wall radiation, proton therapy demonstrated statistically significant reductions in mean dose exposure to the heart, lungs, and left anterior descending artery (LAD) compared to photon therapy. Analysis using the Wilcoxon signed-rank test revealed a W-value of 0 for heart dose, corresponding to a significant reduction with a mean difference of -792.45 cGy. The sum of positive ranks was 0, while the sum of negative ranks was 21, with a computed Z-value of -2.2014. Similarly, proton therapy significantly reduced lung exposure, showing a W-value of 0, a mean difference of -820.4 cGy, a sum of negative ranks totaling 21, and a Z-value of -2.2014. For the LAD, proton therapy also demonstrated a statistically significant reduction in mean dose exposure, with a W-value of 0, a mean difference of -1117.01 cGy, a sum of negative ranks totaling 28, and a Z-value of -2.3664. Although the sample sizes (N = 6 for heart and lungs, N = 7 for LAD) limited the ability to precisely calculate p-values due to insufficient distribution normality, the observed W-values matched or exceeded the critical values for statistical significance at p < 0.05. These results confirm that proton therapy significantly lowers mean doses to the heart, lungs, and LAD compared to photon therapy in this subset of patients.

## Discussion

This study shows a striking difference between IMPT and VMAT in OAR sparing to the lungs, heart, and LAD in sBBC patients. VMAT was chosen as the comparison radiotherapy to IMPT because previous research had already shown that VMAT tends to spare normal tissue more effectively than other radiotherapy techniques [8]. Despite this, the study found an overwhelmingly significant difference in normal tissue exposure when comparing IMPT to VMAT.

Three similar studies have shown findings consistent with our research. Sun et al. looked at 11 patients and found that IMPT plans decreased average target coverage and spared the heart, lungs, LAD, and left ventricle more than VMAT [24]. Similarly, Garda et al. examined another cohort of 11 patients and found that IMPT led to lowered mean doses to the heart and lungs, as well as reduced dosing to the LAD and right coronary artery [23]. Stick et al., in a study of 24 patients, found that proton therapy allows for a significantly lower dose of radiation to the heart and lung and reduces the risk of coronary artery events, lung and breast fibrosis, radiation pneumonitis, and secondary lung cancer. In 17 of the 24 photon plans, the primary clinical target volume of V95% and/or nodal clinical target volume of V90% did not go below 95% in any of the proton plans [25]. The Stick et al. study did not look at LAD dosing, while the former two did and showed similar findings to our present study [23–25]. Overall, the findings of this study are consistent with previous research and contribute to the limited body of literature on radiation dosimetry in patients with sBBC.

While IMPT has clear benefits, access remains a challenge for many patients. IMPT is more expensive than other forms of radiotherapy and is often not approved by insurance, causing delays in care [27–29]. National studies have shown approval rates for IMPT in breast cancer to be as low as 50%, and even after prior authorization, many will still be denied [30]. Despite the expansion of proton therapy, significant patient populations still lack access to this treatment modality. A national study with 871 proton patients and 723,621 non-proton patients examined demographic factors influencing access to proton therapy and found that it is generally associated with Caucasian patients, higher median incomes (over $63,000 per year), residents of southern and western regions of the USA, and those living in metropolitan areas [31]. Establishing the long-term benefits of IMPT in normal tissue sparing and complication prevention is vital for advocating increased insurance coverage, particularly for sBBC patients who are at higher risk from bilateral chest radiation.

Previous research has compared the risks of radiation to its survival benefits in breast cancer. A comprehensive study conducted over 10 years found that 14.17% of mortality was attributed to breast cancer while 5.39% was due to cardiovascular disease [32]. Therefore, minimizing radiation exposure to normal tissue is crucial for improving outcomes in breast cancer patients, especially those with sBBC receiving radiation on both sides. This study’s findings suggest that IMPT shows great promise in enhancing outcomes for sBBC patients.

Long-term follow-up is needed to assess long-term cardiovascular disease outcomes and other adverse effects of radiation, such as secondary malignancy. Prior studies have shown that proton therapy may lead to fewer predicted secondary malignancies in thymic radiation than photon therapy [32]. Further research is needed to confirm this relationship with chest irradiation in sBBC. An improved overall survival rate could incentivize insurance companies to authorize IMPT as a standard of care for sBBC.

Importantly, the significant reduction in mean dose exposure to critical structures such as the heart, lungs, and LAD observed specifically in direct chest wall radiation further underscores the clinical benefit of proton therapy. Despite this clear advantage, current insurance policies frequently restrict proton therapy approvals to selective indications, such as pediatric cancers, central nervous system tumors, unresectable lung cancers, hepatocellular carcinoma, mediastinal lymphoma, and re-irradiation, but notably omit direct chest wall irradiation for breast cancer. Even as most major insurers are expanding coverage through prior-authorization processes and exception appeals treatments involving direct chest wall radiation typically fall outside established approval pathways. The substantial reduction in dose demonstrated by our study highlights an unmet clinical need for reconsideration of these coverage criteria. Expanding insurance coverage based on robust dosimetric evidence, such as provided by this study, could significantly improve patient access to proton therapy and reduce morbidity associated with bilateral chest wall irradiation.

One limitation of the present study is the small sample size, which is also a challenge noted in the other three studies mentioned. This limitation is likely due to the fact that patients with sBBC represent only about 3-4% of all breast cancer cases nationwide, coupled with the limited number of proton therapy centers across the United States [2]. Since the patients in the study were selected for planning both IMPT and VMAT to enable accurate comparisons, the sample size was consequently restricted. Future studies with larger sample sizes are essential to fully elucidate the relationship between IMPT and OAR sparing. A multi-institutional study would significantly help in increasing the sample size.

## Conclusion

In the challenging case of sBBC, IMPT allows for conformal treatment of the bilateral breasts/chest wall with regional nodes while dramatically sparing the heart, LAD, and lungs. In contrast, VMAT provides plans that can lead to severe lung toxicity and cardiovascular disease. This stark difference in OAR sparing, even in a small sample, argues that IMPT should be incorporated in the treatment of sBBC, especially in breast cancer patients receiving radiation directly to their chest wall.

## Data Availability

All relevant data are within the manuscript and its Supporting Information files.

## Funding

No funding was received by any author during the creation of this manuscript.

## Disclosures

None.

## Acknowledgements

The data used in this article have been included and made available in compliance with the principles incorporated by the PLOS One Journal. All data analysis has been made available in Figures 1-7 and Table 1. I, Noah Khosrowzadeh, had full access to all the data in the study and took responsibility for the integrity of the data and the accuracy of the data analysis. Noah Khosrowzadeh, was the lead author of this manuscript. Kyle Chambers, Aren Singh Saini, Kayla Samimi, Matthew Gompels, Dr. Washington, and Dr. Meshman all assisted in supervision and editing of this document.

## Notes

### Competing Interest Statement

The authors have declared no competing interest.

### Funding Statement

The author(s) received no specific funding for this work.

### Author Declarations

This study was approved by the Institutional Review Board (IRB) at the University of Miami Miller School of Medicine (IRB approval number: # 20090856).

## References

1. Bray F, Laversanne M, Sung H, Ferlay J, Siegel RL, Soerjomataram I, Jemal A. Global cancer statistics 2022: GLOBOCAN estimates of incidence and mortality worldwide for 36 cancers in 185 countries. CA Cancer J Clin. 2024;74(3):229–63. Epub 20240404. doi: 10.3322/caac.21834. PubMed PMID: 38572751.

2. Tada K. Bilateral Breast Cancer. Am Surg. 2025;91(5>):854-8. Epub 20250221. doi: 10.1177/00031348251323713. PubMed PMID: 39982251.

3. Sakai T, Ozkurt E, DeSantis S, Wong SM, Rosenbaum L, Zheng H, Golshan M. National trends of synchronous bilateral breast cancer incidence in the United States. Breast Cancer Res Treat. 2019;178(1):161–7. Epub 20190719. doi: 10.1007/s10549-019-05363-0. PubMed PMID: 31325072.

4. Jobsen JJ, van der Palen J, Ong F, Riemersma S, Struikmans H. Bilateral breast cancer, synchronous and metachronous; differences and outcome. Breast Cancer Res Treat. 2015;153(2):277–83. Epub 20150813. doi: 10.1007/s10549-015-3538-5. PubMed PMID: 26268697.

5. Schwentner L, Wolters R, Wischnewsky M, Kreienberg R, Wockel A. Survival of patients with bilateral versus unilateral breast cancer and impact of guideline adherent adjuvant treatment: a multi-centre cohort study of 5292 patients. Breast. 2012;21(2):171–7. Epub 20110925. doi: 10.1016/j.breast.2011.09.007. PubMed PMID: 21945313.

6. Verkooijen HM, Chatelain V, Fioretta G, Vlastos G, Rapiti E, Sappino AP, et al. Survival after bilateral breast cancer: results from a population-based study. Breast Cancer Res Treat. 2007;105(3):347–57. Epub 20061221. doi: 10.1007/s10549-006-9455-x. PubMed PMID: 17186359.

7. Gollamudi SV, Gelman RS, Peiro G, Schneider LJ, Schnitt SJ, Recht A, et al. Breast-conserving therapy for stage I-II synchronous bilateral breast carcinoma. Cancer. 1997;79(7):1362–9. doi: 10.1002/(sici)1097-0142(19970401)79:7<1362::aid-cncr14>3.0.co;2-y. PubMed PMID: 9083159.

8. Salim N, Popodko A, Tumanova K, Stolbovoy A, Lagkueva I, Ragimov V. Cardiac dose in the treatment of synchronous bilateral breast cancer patients between three different radiotherapy techniques (VMAT, IMRT, and 3D CRT). Discov Oncol. 2023;14(1):29. Epub 20230302. doi: 10.1007/s12672-023-00636-z. PubMed PMID: 36862205; PubMed Central PMCID: PMCPMC9981832.

9. Ares C, Khan S, Macartain AM, Heuberger J, Goitein G, Gruber G, et al. Postoperative proton radiotherapy for localized and locoregional breast cancer: potential for clinically relevant improvements? Int J Radiat Oncol Biol Phys. 2010;76(3):685–97. Epub 20090715. doi: 10.1016/j.ijrobp.2009.02.062. PubMed PMID: 19615828.

10. Bouillon K, Haddy N, Delaloge S, Garbay JR, Garsi JP, Brindel P, et al. Long-term cardiovascular mortality after radiotherapy for breast cancer. J Am Coll Cardiol. 2011;57(4):445–52. doi: 10.1016/j.jacc.2010.08.638. PubMed PMID: 21251585.

11. Fiorentino A, Tebano U, Ruggieri R, Ricchetti F, Alongi F. Simultaneous integrated bilateral breast and nodal irradiation with volumetric arc therapy: case report and literature review. Tumori. 2016;102(Suppl. 2). Epub 20161111. doi: 10.5301/tj.5000568. PubMed PMID: 27716877.

12. Acevedo F, Ip T, Orellana M, Martinez G, Gabrielli L, Andia M, et al. Oncological Benefit versus Cardiovascular Risk in Breast Cancer Patients Treated with Modern Radiotherapy. J Clin Med. 2022;11(13). Epub 20220704. doi: 10.3390/jcm11133889. PubMed PMID: 35807180; PubMed Central PMCID: PMCPMC9267636.

13. Gagliardi G, Constine LS, Moiseenko V, Correa C, Pierce LJ, Allen AM, Marks LB. Radiation dose-volume effects in the heart. Int J Radiat Oncol Biol Phys. 2010;76(3 Suppl):S77–85. doi: 10.1016/j.ijrobp.2009.04.093. PubMed PMID: 20171522.

14. Darby SC, Ewertz M, McGale P, Bennet AM, Blom-Goldman U, Bronnum D, et al. Risk of ischemic heart disease in women after radiotherapy for breast cancer. N Engl J Med. 2013;368(11):987–98. doi: 10.1056/NEJMoa1209825. PubMed PMID: 23484825.

15. Jacobse JN, Duane FK, Boekel NB, Schaapveld M, Hauptmann M, Hooning MJ, et al. Radiation Dose-Response for Risk of Myocardial Infarction in Breast Cancer Survivors. Int J Radiat Oncol Biol Phys. 2019;103(3):595–604. Epub 20181029. doi: 10.1016/j.ijrobp.2018.10.025. PubMed PMID: 30385276; PubMed Central PMCID: PMCPMC6361769.

16. Taylor CW, Nisbet A, McGale P, Darby SC. Cardiac exposures in breast cancer radiotherapy: 1950s-1990s. Int J Radiat Oncol Biol Phys. 2007;69(5):1484–95. doi: 10.1016/j.ijrobp.2007.05.034. PubMed PMID: 18035211.

17. Sardaro A, Petruzzelli MF, D’Errico MP, Grimaldi L, Pili G, Portaluri M. Radiation-induced cardiac damage in early left breast cancer patients: risk factors, biological mechanisms, radiobiology, and dosimetric constraints. Radiother Oncol. 2012;103(2):133–42. Epub 20120303. doi: 10.1016/j.radonc.2012.02.008. PubMed PMID: 22391054.

18. Kong FM, Ten Haken R, Eisbruch A, Lawrence TS. Non-small cell lung cancer therapy-related pulmonary toxicity: an update on radiation pneumonitis and fibrosis. Semin Oncol. 2005;32(2 Suppl 3):S42–54. doi: 10.1053/j.seminoncol.2005.03.009. PubMed PMID: 16015535.

19. Mazeron R, Etienne-Mastroianni B, Perol D, Arpin D, Vincent M, Falchero L, et al. Predictive factors of late radiation fibrosis: a prospective study in non-small cell lung cancer. Int J Radiat Oncol Biol Phys. 2010;77(1):38–43. Epub 20100218. doi: 10.1016/j.ijrobp.2009.04.019. PubMed PMID: 20171801.

20. Xu N, Ho MW, Li Z, Morris CG, Mendenhall NP. Can proton therapy improve the therapeutic ratio in breast cancer patients at risk for nodal disease? Am J Clin Oncol. 2014;37(6):568–74. doi: 10.1097/COC.0b013e318280d614. PubMed PMID: 23466577.

21. Bradley JA, Dagan R, Ho MW, Rutenberg M, Morris CG, Li Z, Mendenhall NP. Initial Report of a Prospective Dosimetric and Clinical Feasibility Trial Demonstrates the Potential of Protons to Increase the Therapeutic Ratio in Breast Cancer Compared With Photons. Int J Radiat Oncol Biol Phys. 2016;95(1):411–21. Epub 20150925. doi: 10.1016/j.ijrobp.2015.09.018. PubMed PMID: 26611875.

22. Lin LL, Vennarini S, Dimofte A, Ravanelli D, Shillington K, Batra S, et al. Proton beam versus photon beam dose to the heart and left anterior descending artery for left-sided breast cancer. Acta Oncol. 2015;54(7):1032–9. Epub 20150319. doi: 10.3109/0284186X.2015.1011756. PubMed PMID: 25789715.

23. Garda AE, Hunzeker AE, Michel AK, Fattahi S, Shiraishi S, Remmes NB, et al. Intensity Modulated Proton Therapy for Bilateral Breast or Chest Wall and Comprehensive Nodal Irradiation for Synchronous Bilateral Breast Cancer: Initial Clinical Experience and Dosimetric Comparison. Adv Radiat Oncol. 2022;7(3):100901. Epub 20220124. doi: 10.1016/j.adro.2022.100901. PubMed PMID: 35647397; PubMed Central PMCID: PMCPMC9133394.

24. Sun T, Lin X, Tong Y, Liu X, Pan L, Tao C, et al. Heart and Cardiac Substructure Dose Sparing in Synchronous Bilateral Breast Radiotherapy: A Dosimetric Study of Proton and Photon Radiation Therapy. Front Oncol. 2019;9:1456. Epub 20200110. doi: 10.3389/fonc.2019.01456. PubMed PMID: 31998635; PubMed Central PMCID: PMCPMC6966409.

25. Stick LB, Jensen MF, Bentzen SM, Kamby C, Lundgaard AY, Maraldo MV, et al. Radiation-Induced Toxicity Risks in Photon Versus Proton Therapy for Synchronous Bilateral Breast Cancer. Int J Part Ther. 2022;8(4):1–13. Epub 20211111. doi: 10.14338/IJPT-21-00023.1. PubMed PMID: 35530186; PubMed Central PMCID: PMCPMC9009461.

26. McKenzie E, Razvi Y, Bosnic S, Wronski M, Zhang L, Karam I, et al. Dosimetry and outcomes in patients receiving radiotherapy for synchronous bilateral breast cancers. J Med Imaging Radiat Sci. 2021;52(4):527–43. Epub 20210924. doi: 10.1016/j.jmir.2021.08.013. PubMed PMID: 34580051.

27. Ning MS, Gomez DR, Shah AK, Kim CR, Palmer MB, Thaker NG, et al. The Insurance Approval Process for Proton Radiation Therapy: A Significant Barrier to Patient Care. Int J Radiat Oncol Biol Phys. 2019;104(4):724–33. Epub 20181214. doi: 10.1016/j.ijrobp.2018.12.019. PubMed PMID: 30557675.

28. Gupta A, Khan AJ, Goyal S, Millevoi R, Elsebai N, Jabbour SK, et al. Insurance Approval for Proton Beam Therapy and its Impact on Delays in Treatment. Int J Radiat Oncol Biol Phys. 2019;104(4):714–23. Epub 20181214. doi: 10.1016/j.ijrobp.2018.12.021. PubMed PMID: 30557673; PubMed Central PMCID: PMCPMC10915745.

29. Mendenhall WM, Smith S, Morris CG, Bradley JA, Mailhot Vega RB, McIntyre K, et al. Insurance Coverage for Adjuvant Proton Therapy in the Definitive Treatment of Breast Cancer. Int J Part Ther. 2019;6(2):26–30. Epub 20191011. doi: 10.14338/IJPT-19-00070.1. PubMed PMID: 31998818; PubMed Central PMCID: PMCPMC6986398.

30. Brooks ED, Sio TT, Ning MS, Morris CG, Mendenhall NP, Turner M, et al. Independent Review Organization and Proton Therapy: Multistate Analysis and Legal Procedural Strategies. Int J Part Ther. 2025;15:100741. Epub 20250217. doi: 10.1016/j.ijpt.2025.100741. PubMed PMID: 40084161; PubMed Central PMCID: PMCPMC11905844.

31. Chowdhary M, Lee A, Gao S, Wang D, Barry PN, Diaz R, et al. Is Proton Therapy a "Pro" for Breast Cancer? A Comparison of Proton vs. Non-proton Radiotherapy Using the National Cancer Database. Front Oncol. 2018;8:678. Epub 20190114. doi: 10.3389/fonc.2018.00678. PubMed PMID: 30693271; PubMed Central PMCID: PMCPMC6339938.

32. Fu J, Wu L, Jiang M, Li D, Jiang T, Fu W, et al. Real-world impact of non-breast cancer-specific death on overall survival in resectable breast cancer. Cancer. 2017;123(13):2432–43. Epub 20170307. doi: 10.1002/cncr.30617. PubMed PMID: 28267199.

